# Outcomes of children with serious traumatic brain injury treated in pediatric vs. adult departments

**DOI:** 10.1101/2024.10.22.24315939

**Authors:** Nora Bruns, Rayan Hojeij, Pia Brensing, Michael Nonnemacher, Philipp Dammann, Marcel Dudda, Ursula Felderhoff-Müser, Andreas Stang, Christian Dohna-Schwake

## Abstract

The impact of treatment in a non-pediatric department on mortality and outcomes of children with traumatic brain injury (TBI) is unknown. This study aimed to quantify the impact of the treating department (pediatric (PD) or adult (AD)) on in-hospital case fatality and functional short-term outcomes in children with serious TBI who survived the initial 12 hours after hospital admission.

A Nationwide retrospective cohort study was conducted from a dataset that comprises all discharges from German hospitals from 2016 to 2021. Disease and procedural codes were used to retrieve clinical and outcome information. Hierarchical multilevel logistic regression modeling was performed to quantify the effect of the treating department on the outcomes of interest. Eligibility criteria were age < 18 years and hospital admission with serious TBI.

The main outcome was in-hospital death and secondary outcomes were pediatric complex chronic conditions category (PCCC) ≥ 2 in survivors, composite outcome (death or PCCC ≥ 2), and adjusted mean PCCC in survivors.

Of 13,492,528 pediatric cases, 12,275 were included. The adjusted odds ratio (OR) for death was 3.00 (95 % confidence interval 1.93–4.68) for children treated in ADs compared to PDs. The OR for PCCC ≥ 2 was 0.93 (0.78–1.12) and 1.04 (0.87–1.25) for the composite outcome.

Adjusted average PCCC were 0.40 (0.37–0.44) in ADs and 0.44 (0.42–0.46) in PDs.

This comprehensive nationwide study found increased odds for in-hospital death but similar functional outcomes at discharge among surviving children with serious TBI who were treated outside of PDs.

## Introduction

Traumatic brain injury (TBI) is among the most important causes of acquired morbidity and mortality in children and adolescents worldwide [1, 2]. In Germany, approximately 2,500 children experience a traumatic intracranial injury each year, with approximately 600 requiring a neurosurgical intervention [3]. About 90 children with TBI die annually in German hospitals [3], with an unknown number of pre-hospital deaths.

Besides neurosurgical interventions to evacuate mass lesions and relieve elevated intracranial pressure, the main therapeutic strategy in patients with TBI is to prevent secondary damage through normalization of physiological parameters. From the accident site, in the shock room and later throughout the ICU treatment, optimal medical care is achieved by adjustment and titration of, e.g., oxygenation, carbon dioxide levels, blood pressure, body temperature, blood glucose levels, intracranial pressure, and cerebral perfusion pressure [4-6]. Specialized equipment tailored to the child’s size, profound knowledge on age-specific physiological demands, and practical experience in treating critically ill children are indispensable in this context. Logically, these requirements can only be partially demanded from professionals and departments that do not routinely care for children.

In Germany, there is no requirement specifying whether critically ill or injured children should be admitted to an adult (AD) or pediatric department (PD). From the age of 14 years on, hospitals can use adult procedural codes for “complex intensive care treatment” for billing purposes (www.dimdi.de), suggesting that adolescents from this age onward can be treated by adult physicians. In practice, much younger children and even infants, are treated in AICUs in Germany.

However, physiological demands and pathophysiology evolve rapidly during childhood and clearly differ from those of adults, which becomes particularly evident in the pathophysiological processes following traumatic brain injury [7]. Data from the United States indicate increased mortality in children with polytrauma who are not treated in specialized pediatric trauma centers [8-10]. In Germany, the care of polytraumatized children under 12 years of age is regulated in the “White Book Polytrauma Care” and a national guideline on pediatric polytrauma care to ensure emergency room treatment corresponds to children’s physiological needs [11, 12]. Yet, these recommendations do not apply for the subsequent intensive care treatment.

The aim of this study was to quantify the impact of the treating department (pediatric or adult) on in-hospital case fatality and functional status at hospital discharge in children with serious TBI who survived the initial stabilization in the shock room.

## Methods

The German hospital dataset (GHD) is a nationwide dataset comprising all hospitalizations in public hospitals in the country. Since 2004, German hospitals receive compensation based on diagnosis related groups (DRG). As per §21 KHEntgG, it is mandated by law that German hospitals share data on all hospital admissions with the Hospital Remuneration System

(InEK). After passing plausibility checks, this data is anonymized and forwarded to the Federal Statistical Office. Given that the provision of hospitalization data is obligatory for reimbursement, hospitals have a strong incentive to provide comprehensive data. Detailed information on the structure of the DRG dataset is available at the Federal Statistical Office and further details on the process of data access at https://www.forschungsdatenzentrum.de/en/health/drg.

For this nationwide retrospective cohort study, we extracted TBI cases from 13,492,528 hospital admissions of individuals aged < 18 years between 2016 and 2022 from the GHD.

### Case selection

Cases with traumatic brain injury (ICD-10 code: S06) as primary or secondary discharge diagnosis were identified via codes of the International Classification of Diseases, 10^th^ Edition, German Modification (ICD-10-GM). The severity of the head injury was calculated based on the abbreviated injury scale (AIS) [13, 14] and cases with AIS head ≥ 3 were selected for analyses. Cases with fatal prognosis that deceased within 12 hours of admission were not analyzed (detailed explanation in section “treating department”). Transferred patients were included in descriptive statistics but excluded from regression analyses to avoid analysis of duplicate cases.

### Data extraction

Primary and secondary diagnoses and medical procedures were extracted via ICD-10 codes and codes for surgeries and procedures (Operations-und Prozedurenschlüssel, OPS) to assess the course of disease, interventions, organ failure, and clinical outcomes. Organ dysfunction was assessed by creating indicator variables (0 = absent, 1 = present) for several organ dysfunctions that were added to derive a total score (pediatric organ failure (POF) score). Functional status at discharge was assessed using the Pediatric Complex Chronic Conditions (PCCC) Classification [15], with minor modifications due to lack of information on device prescription in the GHD as previously applied [16]. The PCCC was designed to identify conditions in children that are likely to persist for at least one year from ICD-10 codes, with congenital anomalies extensively considered in the score. Higher scores indicate a higher number of organ systems affected by chronic conditions. All extracted and newly calculated items are detailed in Supplementary table S1 and the PCCC calculation in Supplementary table S2.

### Injury severity

Injury severity was measured as previously described using the ICD-ISS map to derive the Abbreviated Injury Scale (AIS) and Injury Severity Score (ISS) from ICD-10 codes [14, 17]. Survival risk ratios (SRR) were calculated from on ICD-10 codes as previously described [17]. A lower SRR value indicates higher risk of death. For each case, the single worst injury (= lowest SRR value) was extracted from all trauma-related ICD codes. The multiplicative injury severity score, which is designed to account for multiple injuries, was calculated by multiplying the assigned SRR values of all trauma-related ICD codes of each case. These scores were used to categorize and describe injury severity (AIS head, maximum AIS, ISS, single worst injury, multiplicative injury severity score) and adjust for injury severity in regression analyses (single worst injury).

### Primary and secondary endpoints

The unit of analysis was hospital admission with TBI. The primary endpoint was in-hospital death. Secondary endpoints were a PCCC score ≥ 2 and the adjusted mean PCCC score per group. A PCCC score ≥ 2 was chosen as a clinically relevant outcome to avoid overestimating the proportion of poor outcomes, because minor congenital anomalies can cause the PCCC to be 1 and because the rehabilitative potential in the early phase after TBI is high.

### Treating department

In Germany, trauma centers are not divided into pediatric, adult or mixed trauma centers. Even though certifications for becoming a renowned pediatric trauma center can be requested, all trauma centers are allowed to treat injured children. Because we were interested in the impact of management in the intensive care unit among shock room survivors, we extracted information on the department (pediatric versus adult) in which the patient was treated 12 hours after hospital admission. Cases treated in a department with a pediatric department code (general pediatrics, neonatology, pediatric cardiology, pediatric surgery, and intensive care medicine with specialty in pediatrics) were defined as treated in a pediatric department (PD). All cases treated outside of one of these types of departments were considered as treated in adult departments (AD).

In case a patient died within the first 12 hours of admission, the discharging department was considered as the treating department. Because emergency departments and shock rooms frequently use adult department codes and to avoid bias from unpreventable early deaths, cases that died within 12 hours of hospital admission were excluded from regression analyses.

### Missing Data

No data were missing on age and primary diagnoses. Missingness of secondary diagnoses and procedures could not be determined because it was impossible to differentiate whether the diagnosis was truly absent or not coded. We assumed that well-reimbursed diagnoses and procedures were coded comprehensively, and therefore focused data extraction on these codes.

### Statistical analyses

Categorical variables are presented as counts and continuous variables as means (if normally distributed) or medians (if skewed).

We performed hierarchical logistic and linear regression for the primary and secondary endpoints, as appropriate. Because patients treated in the same center were not independent, we accounted for clustering within centers by hierarchical modeling using the treating hospital (identified via the institutional identifier) as a random effect [9, 18]. We identified minimally sufficient adjustment sets for regression analyses using causal diagrams based on the theory of directed acyclic graphs [19-21] as recommended for causal inference in pediatric and critical care research [22, 23]. According to the DAG, we adjusted for age and injury severity (Figure 1) and added the treatment center as random effect as described above. To adjust for injury severity, we used the survival risk ratio from the single worst injury, because this measure has previously been shown to be the superior adjustment measure in general and pediatric trauma patients in administrative datasets, including the GHD [17, 24, 25].

**Figure 1.**
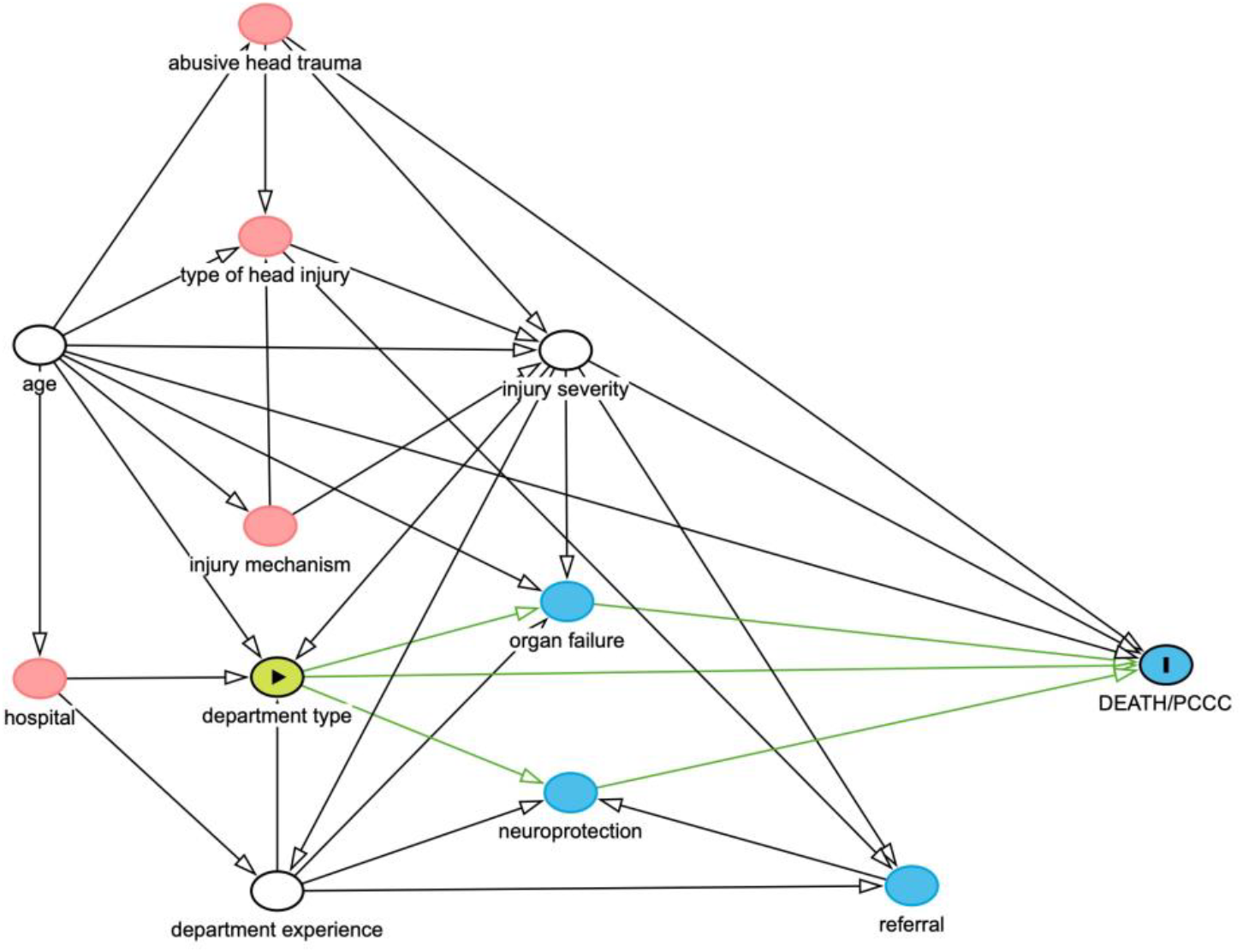
Directed acyclic graph to identify the minimally sufficient adjustment set for multivariable analyses. Green circle: exposure of interest; blue circle with “I”: outcome; white circles: covariates in regression model. Minimally sufficient adjustment set: age, injury severity, department.

### Software

All calculations were carried out using SAS release 9.4 and SAS Enterprise Guide 7.1 (SAS Institute, Cary, North Carolina, USA).

## Results

Of 13,492,528 pediatric cases in the GHD from 2016 - 2022, 577,649 had TBI as discharge diagnose and 14,222 had AIS head ≥ 3 (Figure 2). 1,738 (12.5 %) cases were transferred from the initial hospital and therefore excluded from analyses. Of the remaining 12,439 cases, 164 (1.3 %) died within the first 12 hours of admission (53 (32.3 %) in PDs and 111 (67.7 %) in ADs) (Table 1).

**Table 1.** Baseline characteristics of deceased patients by department, including early deaths within 12 hours after admission.

|  | Early death |  | Late death |  |
| --- | --- | --- | --- | --- |
|  | Pediatric department (n = 53) | Adult department (n = 111) | Pediatric department (n = 171) | Adult department (n = 156) |
| Age | 3 (0 – 8) | 13 (4 – 17) | 2 (0 – 8) | 16 (11 – 17) |
| AIS head | 3 (3 – 3) | 3 (3 – 3) | 3 (3 – 3) | 3 (3 – 3) |
| ISS score | 13 (9 – 22) | 20.5 (14 – 27) | 13 (9 – 19) | 18 (10 – 22) |
| Maximum AIS | 3 (3 – 3) | 3 (3 – 3) | 3 (3 – 3) | 3 (3 – 3) |
| Single worst injury (SRR) | 0.07 (0.07 – 0.50) | 0.38 (0.07 – 0.57) | 0.48 (0.07 – 0.57) | 0.55 (0.38 – 0.62) |
| Multiplicative injury score | 0.03 (0.00 – 0.11) | 0.02 (0.00 – 0.20) | 0.03 (0.00 – 0.11) | 0.04 (0.01 – 0.12) |
| Time to death (hours) | 3.6 (1.2 – 6.1) | 1.9 (0.8 – 3.9) | 89.6 (45.4 – 168.3) | 79.3 (33.2 – 155.9) |
| Duration of stay (days) | 1 (1 – 1) | 1 (1 – 1) | 4 (2 – 7) | 3 (1 – 6) |
| Duration of mechanical ventilation (hours) | 4 (3 – 6) | 3 (1 – 6) | 88 (51 – 170) | 81.5 (31 – 151.5) |
| POF sum score | 1 (1 – 1) | 1 (0 – 1) | 1 (1 – 2) | 1 (1 – 2) |

Numbers are presented as median and interquartile range. AIS = abbreviated injury scale, ISS = injury severity score, POF = pediatric organ failure, SRR = survival risk ratio

**Table 2.** Baseline characteristics of children with severe head trauma by treating department.

|  | **Pediatric department**<br>**(n = 9,082)** | **Adult department**<br>**(n = 3,193)** |
| --- | --- | --- |
| Female | 3352 (36.9) | 959 (30.0) |
| Age (years), *median (IQR)* | 4 (0 – 10) | 15 (9 – 17) |
| AIS head, *median (IQR)* | 3 (3 – 3) | 3 (3 – 3) |
| ISS score, *median (IQR)* | 9 (9 – 10) | 10 (9 – 14) |
| Maximum AIS, *median (IQR)* | 3 (3 – 3) | 3 (3 – 3) |
| Single worst injury (SRR) ,<br>*median (IQR)* | 0.96 (0.90 – 0.97) | 0.93 (0.82 – 0.96) |
| Multiplicative injury score,<br>*median (IQR)* | 0.93 (0.78 – 0.95) | 0.86 (0.61 – 0.94) |
| Transferred | 1044 (10.3) | 903 (22.1) |
| Referred early | 552 (6.1) | 349 (10.9) |
| Referred late | 517 (5.7) | 195 (6.1) |
| Cardiopulmonary<br>resuscitation | 94 (1.0) | 66 (2.1) |
| Out of hospital cardiac<br>arrest | 46 (0.5) | 30 (0.9) |
| Coma | 198 (2.2) | 172 (5.4) |
| Intracranial injury |  |  |
| Brain edema | 593 (6.5) | 445 (13.9) |
| Epidural hematoma | 2239 (24.7) | 654 (20.6) |
| Subdural hematoma | 4074 (44.9) | 1225 (38.4) |
| Subarachnoidal<br>hemorrhage | 1015 (11.2) | 627 (19.6) |

Numbers represent n (%) unless otherwise indicated. IQR = interquartile range

**Figure 2.**
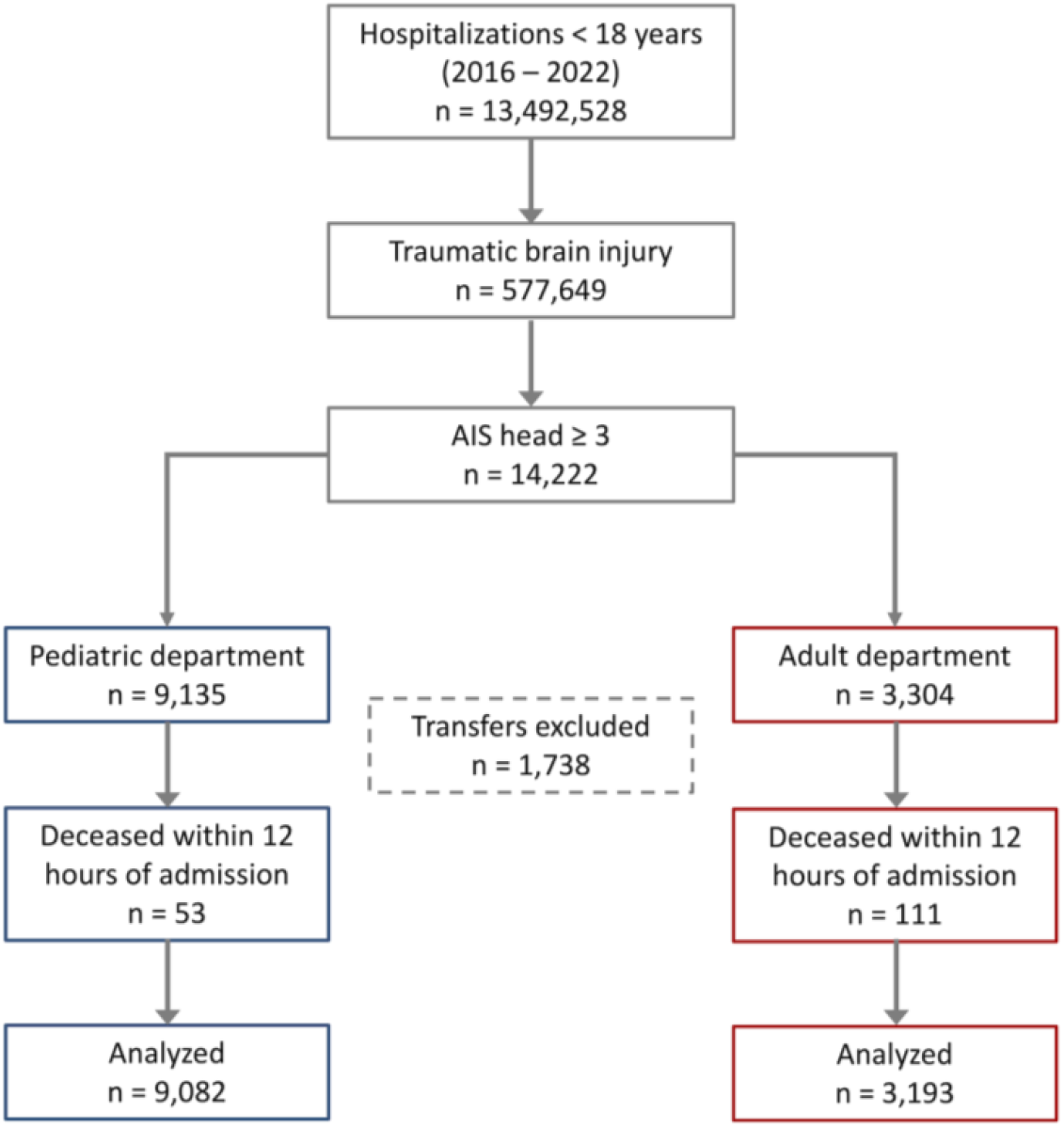
Flow chart of case selection. AIS = Abbreviated Injury Scale

After exclusion of early deaths, 12,275 cases were analyzed, of which 9,082 (74.0 %) were treated in PDs and 3,193 (26.0 %) in ADs (Table 1). The absolute numbers of admissions and deaths were highest in infants and lowest in 8 to 12-year-olds with subsequent rise with increasing age (Figure 3a + b). Case fatality was highest in infants and adolescents. The proportion of admissions to ADs and PDs was inversely correlated with age: the majority of infants was hospitalized in PDs, whereas the proportion of admissions to adult units increased to 83 % in 17-year-olds (Figure 3b).

**Figure 3.**
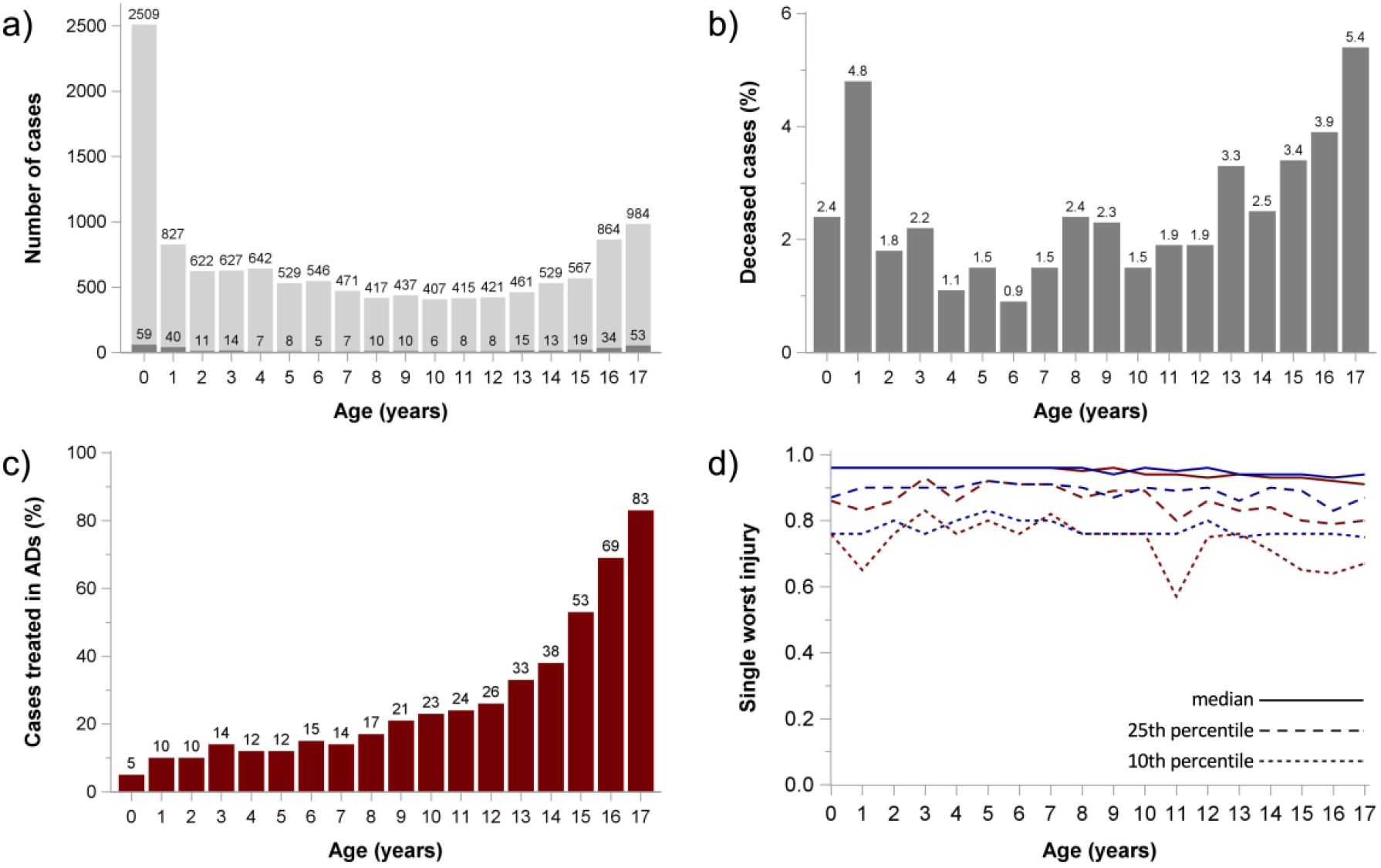
Age-dependency of treated cases a) Number of cases (light grey) and deceased cases (dark grey) b) Case fatality c) Percentage of cases treated in adult departments (AD = adult department) d) Injury severity measured by the single worst injury survival risk ratio (red = adult department, blue = pediatric department)

The overall injury scores showed more severe injuries in adolescents than in younger children with differences between ADs and PDs only in the percentiles that reflect most severe injuries (10^th^ or 90^th^ percentile, respectively, depending on score) (Table 3, figure 3). Crude odds ratios of children treated in ADs versus PDs were 3.24 (95 % CI 2.60 – 4.04) for death, 1.35 (1.20 – 1.53) for PCCC ≥ 2, and 1.52 (1.34 – 1.71) for the composite outcome (death or PCCC ≥ 2). The mean unadjusted PCCC values in survivors were 0.52 (0.48 – 0.55) in ADs and 0.41 (0.40 – 0.43) in PDs.

**Table 3.** Outcomes of children with severe head trauma by treating department.

|  | **Pediatric department**<br>**(n = 9,082)** | **Adult department**<br>**(n = 3,193)** |
| --- | --- | --- |
| Duration of stay (days),<br>*median (IQR)* | 6 (3 – 10) | 6 (3 – 14) |
| Duration of mechanical<br>ventilation (hours), *median*<br>*(IQR)* | 47 (7 – 146) | 80 (17 – 221) |
| PCCC in survivors, *median*<br>*(IQR)* | 0 (0 – 1) | 0 (0 – 1) |
| PCCC ≥ 1 | 2447 (26.9) | 1009 (31.6) |
| PCCC ≥ 2 | 945 (10.4) | 433 (13.6) |
| Deceased | 156 (1.7) | 171 (5.4) |
| Deceased or PCCC ≥ 1 | 2470 (27.2) | 1045 (32.7) |
| Deceased or PCCC ≥ 2 | 1030 (11.3) | 521 (16.3) |
| Seizures | 542 (6.0) | 181 (5.7) |
| Epileptic state | 123 (1.4) | 9 (0.3) |
| POF sum score, *median*<br>*(IQR)* | 0 (0 – 0) | 0 (0 – 1) |
| POF |  |  |
| Cardiological | 71 (0.8) | 65 (2.0) |
| Pulmonary | 1350 (14.9) | 711 (22.3) |
| Coagulation | 38 (0.4) | 29 (1.2) |
| Renal | 35 (0.4) | 38 (1.2) |
| Hepatic | 41 (0.5) | 14 (0.4) |
| Central nervous system | 376 (4.1) | 99 (3.1) |
| Extracorporeal membrane<br>oxygenation | 6 (0.07) | 15 (0.47) |
| Neurosurgery | 1978 (21.8) | 918 (28.8) |
| Evacuation of hematoma | 500 (5.5) | 203 (6.4) |
| Invasive ICP monitoring | 377 (2.5) | 301 (4.2) |
| Extraventricular drain | 240 (2.6) | 102 (3.2) |
| Decompressive<br>craniectomy | 310 (3.4) | 239 (7.5) |
| Other surgery | 1456 (16.0) | 1104 (34.6) |

Numbers represent n (%) unless otherwise indicated. ICP = intracranial pressure IQR = interquartile range, PCCC = pediatric complex chronic conditions score, POF = pediatric organ failure

Hierarchical multilevel regression analyses yielded an odds ratio (OR) for death of 3.00 (95 % CI 1.93– 4.68) if children were treated in ADs compared to PDs. The OR for PCCC ≥ 2 was 0.93 (0.78 – 1.12) in surviving children treated in ADs compared to PDs and 1.04 (0.87 – 1.25) for the composite outcome. Adjusted average PCCC in survivors were 0.40 (0.37 – 0.44) in Ads and 0.44 (0.42 – 0.46) in PDs.

## Discussion

This study from a comprehensive nationwide hospital database found that about 25 % of seriously head-injured children in Germany that survived the initial 12 hours after admission were treated in adult departments. The proportion was age-dependent and increased with age. Cases treated in ADs had slightly more injuries with slightly higher overall injury severity scores. Adjusted odds ratios were higher for death in AD cases but functional outcomes in survivors and the composite outcome were similar between ADs and PDs.

Previous studies have reported increased odds for death in polytraumatized pediatric patients when treated outside dedicated pediatric trauma centers [8-10]. Our study found clearly increased odds for death in children that survived the initial stabilization and were treated in ADs instead of PDs after correcting for age and injury severity, aligning with these reports. Regarding functional outcomes, however, we found only a slight advantage of PDs with respect to the composite outcome death/poor neurological function.

Pre-hospital determinants of outcome and mortality are well understood and include age, injury severity, and the injury pattern [26, 27]. The duration of pre-hospital management is less important than its quality and longer duration of transport to a high-level care center is compensated by faster processes in the emergency department [28, 29]. In children, helicopter emergency transport to high-level care center translates into lower mortality and better outcomes [30, 31]. In contrast, in-hospital determinants of outcome are more complex and less well-understood.

For example, a center’s pediatric trauma patient volume is the only factor that positively influences outcomes after severe pediatric TBI, while adult general trauma and adult TBI patient volumes have no impact [32]. As TBI is frequently the leading injury and the leading cause of death in pediatric polytrauma patients, making optimum care to prevent secondary damage and preserve neurological function crucial [33-35]. Neuroprotection in manifest or suspected TBI must be initiated at the accident site and is to be continued for several days, depending on the type and severity of the primary injury, the degree of secondary damage, and clinical course [4-6, 36]. It extends well into the ICU stay, requiring careful titration of therapeutic measures to achieve physiological homeostasis and prevent secondary damage, along with advanced neuromonitoring techniques [4-6, 36, 37]. Even for experienced intensive care providers, this can be challenging and is further complicated by children’s age-specific physiological characteristics and demands.

At the same time, management decisions can affect case fatality and functional outcomes in critically ill TBI patients: In the RESCUEicp trial lethality was lower in patients receiving decompressive craniectomy compared to conservative management at the cost of severe disability [38]. Given this association, different survival rates may also reflect different decision-making processes regarding interventions and/or withdrawal of care between adult and pediatric departments. This possibility is supported by our finding that the early-deceased children died later in PDs in spite of being more severely injured, suggesting that active life support was withdrawn later than in ADs. Outweighing a child’s death versus potential life-long severe disability is a stressful process that may be influenced by past experiences of the treatment team and the parents’ will [39]. It seems plausible that differences between adult and pediatric caregivers exist.

Apart from lethality, this study found similar functional outcomes between ADs and PDs with respect to the adjusted PCCC score and slightly lower odds for poor functional outcome. It may be assumed that the lower odds for poor functional outcome were achieved by the higher lethality in ADs and consequently depletion of susceptible individuals. For that reason, we calculated a composite outcome that comprised death and poor functional outcome. The adjusted odds for this composite outcome were comparable between both department types. In this context, it is important to take into account the long recovery potential for motor and cognitive functioning after severe TBI. The most rapid recovery occurs during the first year but improvements are observed until several years after the accident [40-42]. As this study could determine only hospital outcomes, longitudinal outcomes should be assessed to deepen the understanding of long-term outcomes in depending on the treating department.

An important limitation of this study is the lack of information on long-term outcomes, because the GHD contains no follow-up data. Because case linking is not allowed in the GHD, patients transferred to other hospitals were excluded from the study to avoid duplicates.

Further, some patients may have been transferred to another department within the hospital during their stay – however, we focused on an early timepoint after admission because the initial phase post-injury is the most dynamic. Assessing neurological functioning from ICD-10 and OPS codes is challenging. We chose the PCCC because it accounts for chronic conditions that are likely to last for one year or longer and also accounts for device-dependency like tracheostomy or percutaneous endoscopic gastrostomy/jejunostomy tubes. However, it must be pointed out that congenital conditions like malformations also raise the PCCC score.

Because underlying chronic conditions or serious malformations are rare in pediatric trauma patients, the PCCC seems nonetheless a suitable method for functional outcome assessment in this specific cohort.

This comprehensive nationwide analysis focused on the association between the treating department and outcomes in severely head-injured children that survived the initial 12 hours after hospital admission. The slightly increased odds for death and the composite outcome in cases that were not treated in pediatric departments imply that best medical care for children is likely provided by dedicated pediatric teams. Prospective studies should be performed and longitudinal datasets exploited to compare long-term morbidity and health care utilization in survivors depending on the treating department. Whether post-stabilization transfer to a pediatric ICU from an external hospital is advisable cannot be deduced from the present study, but should also be examined in the future.

## Conclusion

This study provides evidence that treatment of children with serious TBI in a pediatric department is associated with lower case fatality and comparable neurological short-term function in survivors. Taking into account the recovery potential after severe TBI, the impact of the treating department on long-term outcomes should be investigated by analyzing long-term trajectories, either from longitudinal administrative health care data or prospective studies.

## Declarations

### Data availability

The analyzed dataset remains property of the German research data center and can be accessed by qualified researchers after written inquiry to the corresponding authorities.

### Code availability

The code for statistical analyses will be made available upon reasonable request to any qualified researcher.

### Author contributions

**NB:** conceptualization, data curation, formal analysis, writing – original draft, visualization; **RH:** data curation, writing – review & editing; **PB:** data curation, writing – review & editing; **MN:** data curation, writing – review & editing; **PD:** writing – review & editing; **MD:** writing – review & editing; **UFM:** resources, writing – review & editing; **AS:** conceptualization, resources, software, writing – review & editing; CDS: conceptualization, supervision, writing – review & editing.

## Acknowledgements

N/A

## References

1. Dewan MC, Mummareddy N, Wellons JC, Bonfield CM, (2016) Epidemiology of Global Pediatric Traumatic Brain Injury: Qualitative Review. World neurosurgery 91: 497-509.e491

2. Majdan M, Plancikova D, Brazinova A, Rusnak M, Nieboer D, Feigin V, Maas A, (2016) Epidemiology of traumatic brain injuries in Europe: a cross-sectional analysis. The Lancet Public health 1: e76–e83

3. Bruns N, Trocchi P, Felderhoff-Müser U, Dohna-Schwake C, Stang A, (2021) Hospitalization and Morbidity Rates After Pediatric Traumatic Brain Injury: A Nation-Wide Population-Based Analysis. Front Pediatr 9: 747743

4. Kochanek PM, Tasker RC, Bell MJ, Adelson PD, Carney N, Vavilala MS, Selden NR, Bratton SL, Grant GA, Kissoon N, Reuter-Rice KE, Wainwright MS, (2019) Management of Pediatric Severe Traumatic Brain Injury: 2019 Consensus and Guidelines-Based Algorithm for First and Second Tier Therapies. Pediatric critical care medicine : a journal of the Society of Critical Care Medicine and the World Federation of Pediatric Intensive and Critical Care Societies 20: 269–279

5. Kochanek PM, Tasker RC, Carney N, Totten AM, Adelson PD, Selden NR, Davis-O & apos;Reilly C, Hart EL, Bell MJ, Bratton SL, Grant GA, Kissoon N, Reuter-Rice KE, Vavilala MS, Wainwright MS, (2019) Guidelines for the Management of Pediatric Severe Traumatic Brain Injury, Third Edition: Update of the Brain Trauma Foundation Guidelines, Executive Summary. Pediatric critical care medicine : a journal of the Society of Critical Care Medicine and the World Federation of Pediatric Intensive and Critical Care Societies 20: 280–289

6. Dohna-Schwake C, Rellensmann G, Mauer U, Fitze G, Schmittenbecher P, Baumann F, Sommerfeldt D, Mentzel H-J, Hahn G, Klee D, Becke-Jakob K, Fideler F, Merkenschlager A, Trollmann R, Hoffmann F, Porto L, Wietholt G (2022) AWMF-Leitlinie „Schädelhirntrauma im Kindes-und Jugendalter”. In: Editor (ed)^(eds) Book AWMF-Leitlinie „Schädelhirntrauma im Kindes-und Jugendalter”. AWMF online, City, pp.

7. Giza CC, Mink RB, Madikians A, (2007) Pediatric traumatic brain injury: not just little adults. Curr Opin Crit Care 13: 143–152

8. Potoka DA, Schall LC, Gardner MJ, Stafford PW, Peitzman AB, Ford HR, (2000) Impact of pediatric trauma centers on mortality in a statewide system. J Trauma 49: 237–245

9. Sathya C, Alali AS, Wales PW, Scales DC, Karanicolas PJ, Burd RS, Nance ML, Xiong W, Nathens AB, (2015) Mortality Among Injured Children Treated at Different Trauma Center Types. JAMA Surg 150: 874–881

10. Webman RB, Carter EA, Mittal S, Wang J, Sathya C, Nathens AB, Nance ML, Madigan D, Burd RS, (2016) Association Between Trauma Center Type and Mortality Among Injured Adolescent Patients. JAMA Pediatr 170: 780–786

11. Schmittenbecher P (2020) S2K-Leitlinie „Polytraumaversorgung im Kindesalter”. In: Editor (ed)^(eds) Book S2K-Leitlinie „Polytraumaversorgung im Kindesalter”. City, pp.

12. DGU (2019) Weißbuch Schwerverletztenversorgung. Deutsche Gesellschaft für Unfallchirurgie,

13. Greenspan L, McLellan BA, Greig H, (1985) Abbreviated Injury Scale and Injury Severity Score: a scoring chart. J Trauma 25: 60–64

14. Loftis KL, Price JP, Gillich PJ, Cookman KJ, Brammer AL, St Germain T, Barnes J, Graymire V, Nayduch DA, Read-Allsopp C, Baus K, Stanley PA, Brennan M, (2016) Development of an expert based ICD-9-CM and ICD-10-CM map to AIS 2005 update 2008. Traffic Inj Prev 17 Suppl 1: 1–5

15. Feudtner C, Feinstein JA, Zhong W, Hall M, Dai D, (2014) Pediatric complex chronic conditions classification system version 2: updated for ICD-10 and complex medical technology dependence and transplantation. BMC Pediatrics 14: 199

16. Bruns N, Feddahi N, Hojeij R, Rossi R, Dohna-Schwake C, Stein A, Kobus S, Stang A, Kowall B, Felderhoff-Müser U, Short-term outcomes of asphyxiated neonates depending on requirement for transfer in the first 24 hours of life. Resuscitation

17. Hojeij R, Brensing P, Nonnemacher M, Kowall B, Felderhoff-Müser U, Dudda M, Dohna-Schwake C, Stang A, Bruns N, (2023) Performance of ICD-10-based injury severity scores in pediatric trauma patients using the ICD-AIS map and survival rate ratios. medRxiv: 2023.2012.2004.23299239

18. Austin PC, Goel V, van Walraven C, (2001) An introduction to multilevel regression models. Can J Public Health 92: 150–154

19. Greenland S, Pearl J, Robins JM, (1999) Causal diagrams for epidemiologic research. Epidemiology 10: 37–48

20. Pearl J, (1995) Causal Diagrams for Empirical Research. Biometrika 82: 669–688

21. Textor J, van der Zander B, Gilthorpe MS, Liśkiewicz M, Ellison GTH, (2016) Robust causal inference using directed acyclic graphs: the R package ‘dagitty’. International Journal of Epidemiology 45: 1887–1894

22. Lederer DJ, Bell SC, Branson RD, Chalmers JD, Marshall R, Maslove DM, Ost DE, Punjabi NM, Schatz M, Smyth AR, Stewart PW, Suissa S, Adjei AA, Akdis CA, Azoulay E, Bakker J, Ballas ZK, Bardin PG, Barreiro E, Bellomo R, Bernstein JA, Brusasco V, Buchman TG, Chokroverty S, Collop NA, Crapo JD, Fitzgerald DA, Hale L, Hart N, Herth FJ, Iwashyna TJ, Jenkins G, Kolb M, Marks GB, Mazzone P, Moorman JR, Murphy TM, Noah TL, Reynolds P, Riemann D, Russell RE, Sheikh A, Sotgiu G, Swenson ER, Szczesniak R, Szymusiak R, Teboul JL, Vincent JL, (2019) Control of Confounding and Reporting of Results in Causal Inference Studies. Guidance for Authors from Editors of Respiratory, Sleep, and Critical Care Journals. Ann Am Thorac Soc 16: 22–28

23. Williams TC, Bach CC, Matthiesen NB, Henriksen TB, Gagliardi L, (2018) Directed acyclic graphs: a tool for causal studies in paediatrics. Pediatr Res 84: 487–493

24. Tepas JJ, 3rd, Leaphart CL, Celso BG, Tuten JD, Pieper P, Ramenofsky ML, (2008) Risk stratification simplified: the worst injury predicts mortality for the injured children. J Trauma 65: 1258–1261; discussion 1261-1253

25. Kilgo PD, Osler TM, Meredith W, (2003) The worst injury predicts mortality outcome the best: rethinking the role of multiple injuries in trauma outcome scoring. J Trauma 55: 599–606; discussion 606-597

26. Calland JF, Xin W, Stukenborg GJ, (2013) Effects of leading mortality risk factors among trauma patients vary by age. Journal of Trauma and Acute Care Surgery 75

27. Lefering R, Huber-Wagner S, Nienaber U, Maegele M, Bouillon B, (2014) Update of the trauma risk adjustment model of the TraumaRegister DGU™: the Revised Injury Severity Classification, version II. Critical care (London, England) 18: 476–412

28. Klein K, Lefering R, Jungbluth P, Lendemans S, Hussmann B, (2019) Is Prehospital Time Important for the Treatment of Severely Injured Patients? A Matched-Triplet Analysis of 13,851 Patients from the TraumaRegister DGU®. Biomed Res Int 2019: 5936345

29. Lefering R, Waydhas C, TraumaRegister DGU, (2022) Process times of severely injured patients in the emergency room are associated with patient volume: a registry-based analysis. European Journal of Trauma and Emergency Surgery

30. Bläsius FM, Horst K, Brokmann JC, Lefering R, Andruszkow H, Hildebrand F, TraumaRegister D, (2021) Helicopter Emergency Medical Service and Hospital Treatment Levels Affect Survival in Pediatric Trauma Patients. J Clin Med 10

31. Bruns N, Kamp O, Lange K, Lefering R, Felderhoff-Müser U, Dudda M, Dohna-Schwake C, (2022) Functional Short-Term Outcomes and Mortality in Children with Severe Traumatic Brain Injury: Comparing Decompressive Craniectomy and Medical Management. J Neurotrauma 39: 944–953

32. Kimata AR, Tang OY, Asaad WF, (2021) Generalizability of pediatric major trauma experience to severe pediatric traumatic brain injury at level 1 and 2 trauma centers. Journal of Emergency and Critical Care Medicine 5

33. Schuster A, Klute L, Kerschbaum M, Kunkel J, Schaible J, Straub J, Weber J, Alt V, Popp D, (2024) Injury Pattern and Current Early Clinical Care of Pediatric Polytrauma Comparing Different Age Groups in a Level I Trauma Center. Journal of Clinical Medicine DOI 10.3390/jcm13020639

34. Ringen AH, Baksaas-Aasen K, Skaga NO, Wisborg T, Gaarder C, Naess PA, (2023) Close to zero preventable in-hospital deaths in pediatric trauma patients – An observational study from a major Scandinavian trauma center. Injury 54: 183–188

35. Theodorou CM, Galganski LA, Jurkovich GJ, Farmer DL, Hirose S, Stephenson JT, Trappey AF, (2021) Causes of early mortality in pediatric trauma patients. J Trauma Acute Care Surg 90: 574–581

36. Hawryluk GWJ, Aguilera S, Buki A, Bulger E, Citerio G, Cooper DJ, Arrastia RD, Diringer M, Figaji A, Gao G, Geocadin R, Ghajar J, Harris O, Hoffer A, Hutchinson P, Joseph M, Kitagawa R, Manley G, Mayer S, Menon DK, Meyfroidt G, Michael DB, Oddo M, Okonkwo D, Patel M, Robertson C, Rosenfeld JV, Rubiano AM, Sahuquillo J, Servadei F, Shutter L, Stein D, Stocchetti N, Taccone FS, Timmons S, Tsai E, Ullman JS, Vespa P, Videtta W, Wright DW, Zammit C, Chesnut RM, (2019) A management algorithm for patients with intracranial pressure monitoring: the Seattle International Severe Traumatic Brain Injury Consensus Conference (SIBICC). Intensive Care Med 45: 1783–1794

37. Ko TS, Catennacio E, Shin SS, Stern J, Massey SL, Kilbaugh TJ, Hwang M, (2023) Advanced Neuromonitoring Modalities on the Horizon: Detection and Management of Acute Brain Injury in Children. Neurocrit Care 38: 791–811

38. Hutchinson PJ, Kolias AG, Timofeev IS, Corteen EA, Czosnyka M, Timothy J, Anderson I, Bulters DO, Belli A, Eynon CA, Wadley J, Mendelow AD, Mitchell PM, Wilson MH, Critchley G, Sahuquillo J, Unterberg A, Servadei F, Teasdale GM, Pickard JD, Menon DK, Murray GD, Kirkpatrick PJ, Collaborators RT, (2016) Trial of Decompressive Craniectomy for Traumatic Intracranial Hypertension. The New England journal of medicine 375: 1119–1130

39. Santoro JD, Bennett M, (2018) Ethics of End of Life Decisions in Pediatrics: A Narrative Review of the Roles of Caregivers, Shared Decision-Making, and Patient Centered Values. Behav Sci (Basel) 8

40. Lang SS, Kilbaugh T, Friess S, Sotardi S, Kim CT, Mazandi V, Zhang B, Storm PB, Heuer GG, Tucker A, Ampah SB, Griffis H, Raghupathi R, Huh JW, (2021) Trajectory of Long-Term Outcome in Severe Pediatric Diffuse Axonal Injury: An Exploratory Study. Front Neurol 12: 704576

41. Kolias AG, Adams H, Timofeev IS, Corteen EA, Hossain I, Czosnyka M, Timothy J, Anderson I, Bulters DO, Belli A, Eynon CA, Wadley J, Mendelow AD, Mitchell PM, Wilson MH, Critchley G, Sahuquillo J, Unterberg A, Posti JP, Servadei F, Teasdale GM, Pickard JD, Menon DK, Murray GD, Kirkpatrick PJ, Hutchinson PJ, (2022) Evaluation of Outcomes Among Patients With Traumatic Intracranial Hypertension Treated With Decompressive Craniectomy vs Standard Medical Care at 24 Months: A Secondary Analysis of the RESCUEicp Randomized Clinical Trial. JAMA Neurol 79: 664–671

42. Jaffe KM, Polissar NL, Fay GC, Liao S, (1995) Recovery trends over three years following pediatric traumatic brain injury. Arch Phys Med Rehabil 76: 17–26

