## Supplementary tables for "Outcomes of children with serious traumatic brain injury treated in pediatric vs. adult departments"

**Supplementary Table 1:** Extracted ICD- and OPS-Codes and newly calculated variables

| **Diagnoses** | **ICD-10-Code(s)** |
|---|---|
| Traumatic brain injury | S06 (primary or secondary diagnosis) |
| Intracranial injury | |
| Brain edema | S06.1 |
| Epidural hematoma | S06.4 |
| Subdural hematoma | S06.5 |
| Subarachnoidal hemorrhage | S06.6 |
| Out of hospital cardiac arrest | U69.13 |
| Neurologic complications | |
| Coma | R40.2, S06.72, S06.73 |
| Seizures | G40 |
| Epileptic state | G41 |
| Organ failure | |
| Cardiological (POF_card) | R57 |
| Pulmonary (POF_pulm) | Ventilation = yes |
| Coagulation (POF_coag) | D65.1 |
| Renal (POF_ren) | N17 |
| Hepatic (POF_hep) | K72, R74.0 |
| Central nervous system (POF_CNS) | R40 |

| **Procedures** | **OPS-Codes** |
|---|---|
| Neurosurgery | 5-01, 5-02, 5-03, 5-04, 5-05 |
| Evacuation of hematoma | 5-013.1, 5-013.4, 5-012.2 |
| Invasive ICP monitoring | 5-029.1 |
| Extraventricular drain | 5-022.0 |
| Decompressive craniectomy | 5-021.0, 5-010.1, 5-010.4 |
| Other surgery | 50-6, 5-07, 5-08, 5-09, 5-1, 5-2, 5-3, 5-4, 5-5, 5-6, 5-7, 5-8, 5-9 |
| Dialysis | 8-854, 8-857, 8-858 |
| ECMO (extracorporeal membrane oxygenation) | 8-852 |
| Cardiac or cardiopulmonary resuscitation | I46 |

| **Other variables extracted from the basic data set** | |
|---|---|
| Sex | Included in the DRG data set |
| Length of stay (days) | Included in the DRG data set |
| Ventilation (hours) | Included in the DRG data set |
| Death | Reason for discharge = 7 |
| Transfer | Reason for discharge = 6 or 8 |
| Early referral | Reason for admission = A |
| Late referral | Reason for admission = V |
| Pediatric department | Treating department contains pediatric department code (10xx, 11xx, 12xx, 13xx) |

| **Newly calculated variables** | **Variables/Procedures included [categories]** |
|---|---|
| Ventilation (dichotomous) | Ventilation (hours) not missing [yes, no] |
| Abbreviated injury scale (AIS) head | As described in citation [17] |
| Maximum AIS | |
| Injury severity score | As described in citation [17] |
| Survival risk ratio | As described in citation [17] |
| Single worst injury | As described in citation [17] |
| Multiplicative injury severity score | As described in citation [17] |

| | |
|---|---|
| Resuscitation | Cardiac arrest, cardiac or cardiopulmonary resuscitation [yes, no] |
| Early death | Reason for discharge = 7 and duration of stay ≤ 720 minutes |
| Pediatric organ failure score | Sum of binary scores POF_card, POF_pulm, POF_ren, POF_coag, POF_GI, POF_hep, POF_CNS, Dialysis, ECMO [continuous] |

**eTable 2:** Calculation of the pediatric complex chronic conditions classification

| PCCC subscore | **ICD-10-Code(s)** |
|---|---|
| Neurological (CCC_neuro) | Q00, Q01, Q02, Q03, Q04, Q05, Q06, Q07, G901, F71, F72, F73, E75, F842, G11, G12, G25.3, G31.1, G31.8, G31.9, G32.8, G91.1, G93.8, G93.9, G94, G95.18, G95.88, G90.9, Q85.1, G80, G40, G37.1, G37.2, G37.8, G81, G82, G83.5, G83.9, G93.1, G93.5, R40, I63, G71, G72, G10, G20, G21.1, G21.8, G23.0, G23.1, G23.2, G24.0, G24.8, G25.3, G25.4, G25.5, G25.8, G25.9, T85.0, T85.1, T85.7, Z98.2, Z99 |
| Cardiological (CCC_cardio) | Q20, Q21.2, Q21.3, Q21.4, Q21.8, Q21.9, Q22, Q23, Q24, Q25.1, Q25.2, Q25.3, Q25.5, Q25.6, Q25.7, Q25.8, Q25.9, Q26, Q28.2, Q28.3, Q28.9, I34.0, I34.8, I36.0, I36.8, I37.0, I37.8, I42, I43, I51.5, I44, I45, I47, I48, I49, R00.1, I27, I50.9, I51.7, I51.8, I63.1, I63.2, Z95.1, T82.5, T82.1, T82.0, T82.2, T82.6, T82.7, Z95.0, Z95.2, Z95.3, Z95.8, Z95.9, Z45.01, Z45.02, Z45.09, T86.2, Z94.1 |
| Respiratory (CCC_resp) | Q30, Q31, Q32, Q33, Q34, P28.0, G47.32, I27.8, I43, J84.1, J96, Z90.2, E84, J95.0, J95.8, Z43.0, Z93.0, Z99.0, Z99.1, T86.81, Z94.2 |
| Renal (CCC_renal) | Q60, Q61, Q62, Q63, Q64, N18, Z90.5, Z90.6, G83.4, N31.2, N31.9, T85.71, Z93.5, Z93.6, Z91.1, Z99.2, Z43.5, Z43.6, Z46.6, T86.1, Z94.0 |
| Gastroenterological (CCC_gastro) | Q39.0, Q39.1, Q39.2, Q39.3, Q39.4, Q41, Q42, Q43, Q45, K73, K74, K75, K76.0, K76.1, K76.2, K76.3, K76.5, K76.8, K50, K51, I82.0, K55.1, K56.2, K59.3, Z98.0, Z90.3, Z90.4, Z93.1, Z93.2, Z93.3, Z93.4, Z43.1, Z43.2, Z43.3, Z43.4, T86.4, T86.8, Z94.4, Z94.88 |
| Hematological/immunological (CCC_hema_immu) | D55, D56, D57, D58, D60, D61, D71, D8, D72.0, M30.3, M35.9, D66, D68.2, D69.4, D69.3, D70.0, D70.5, D76.1, D76.2, D76.3, D86.9, B20, B21, B22, B23, B24, M30.0, M31.0, M31.3, M31.4, M31.6, M32.1, M33.9, M34.0, M43.1, M34.9 |
| Metabolic (CCC_metab) | E70.0, E70.2, E70.3, E70.8, E71, E72.0, E72.1, E72.3, E72.4, E72.8, E72.9, E74, E75, E77.0, E77.1, E78, E88.8, E76.0, E76.1, E76.2, E76.3, E85, E78.6, E79.1, E79.8, E80.4, E80.5, E80.6, E80.7, E93.0, E83.1, E83.3, E83.4, D84.1, E88, H49.8, E00.9, E23.0, E23.2, E23.3, E23.7, E24.0, E24.2, E24.3, E24.8, E24.9, E26.8, E25.0, E25.8, E25.9, Z46.8, Z96.4 |
| Genetic malformations (CCC_genet_malform) | Q90.9, Q91.3, Q91.4, Q91.7, Q92.8, Q93, Q95.0, Q96.9, Q97, Q98, Q99.8, Q99.9, E34.3, M41.0, M41.2, M41.39, M41.8, M41.9, M43.3, M96.5, Q72.2, Q75.0, Q75.2, Q75.9, Q76.0, Q76.1, Q76.2, Q76.4, Q76.5, Q76.6, Q77, Q78.0, Q87.1, Q78.2, Q78.3, Q78.4, Q78.8, Q78.9, K44.9, Q79.0, Q79.1, Q79.2, Q79.3, Q79.4, Q79.5, Q79.9, Q81, Q87.1, Q87.2, Q87.3, Q87.4, Q87.8, Q89.7, Q89.9, Q99.2 |
| Malignancy (CCC_malign) | C, D0, E34.0, D37, D38, D39, D4, Q85.0, T86.0, Z94.80, Z84.81 |
| Neonatal (CCC_neo) | P05, P07.0, P07.2, P10.0, P10.1, P10.4, P52.4, P52.8, P11.5, P21.0, P21.9, P96, P25, P27.0, P27.1, P27.8, P91.6, P35.0, P35.1, P35.2, P56.0, P57.0, P57.8, P61.3, P61.4, P77, P83.2, P91.2 |
| Dependency from technical devices (CCC_tech_dep) | T84.01, T84.02, T84.03, T84.04, T84.05, T84.06, T84.08, T84.11, T84.12, T84.18, T84.4, T84.5, T84.6, T84.7, T86.9, T86.0, T86.8, T86.5, T87.0, T87.1, Z99.8, Z93.1, Z93.2, Z93.3, Z93.4, Z43.1, Z43.2, Z43.3, Z43.4, T85.71, Z93.5, Z93.6, Z91.1, Z99.2, Z43.5, Z43.6, Z46.6, J95.0, J95.8, Z43.0, Z93.0, Z99.0, Z99.1, T82.5, T82.1, T82.0, T82.2, T82.6, T82.7, Z95.0, Z95.2, Z95.3, Z95.8, Z95.9, Z45.01, Z45.02, Z45.09, T85.0, T85.1, T85.7, Z98.2, Z99 |
| Transplantation recipient* (CCC_transplant) | Z94, T86 |

| | |
|---|---|
| PCCC score | Each organ specific binary score is set to 1 if one or more conditions (resembled by ICD-10 codes) apply. The PCCC score represents the sum of all binary sores. |

* solid organ or bone marrow transplant
